# Multigene Panel Testing Outcomes in Patients with Uveal Melanoma: Implementation of National Guidelines

**DOI:** 10.64898/2025.11.26.25341005

**Authors:** Lindsey Byrne, Isabella Gray, Kaylee Ramsey, Joseph McElroy, Emma Schreiner, Jenna Wolfe, Olivia B. Taylor, Frederick Davidorf, Colleen M. Cebulla, Mohamed H. Abdel-Rahman

**Author notes:** Mohamed H Abdel-Rahman, MD, PhD; Department of Ophthalmology and Visual Sciences; The Ohio State University Wexner Medical Center; 400 W 12^th^ Ave, 225 Wiseman Hall, Columbus, Ohio 43210. **Data Availability Statement:** The data presented in this study are available on request from the corresponding author. **Ethics approval:** This work was done under research protocols approved by the Institutional Review Board of The Ohio State University.

## Abstract

This study aimed to evaluate the outcome of germline clinical genetic testing in uveal melanoma (UM) patients who met the National Comprehensive Cancer Network (NCCN) guidelines for genetic testing. A retrospective chart review was conducted on UM patients seen in The Ohio State University Cancer Genetics Clinic between 5/1/2021-9/19/2025. Seventy individuals underwent clinical genetic testing, primarily via large multi-gene panels. Ten UM patients, including two related individuals, had pathogenic or likely pathogenic (P/LP) variants in known cancer genes (*BAP1, BRCA1, BRCA2, MBD4, MUTYH, POT1, XRCC2)*. Among unrelated individuals, the positive rate was12.9% (8/70). Excluding carrier genes, the rate was10% (7/70) Eight patients would have been missed if only tested for *BAP1* per ASCO 2024 recommendations. There was no association between tumor size, stage and germline P/LP variants. In summary, NCCN guidelines are useful in the prioritization of UM patients for genetic testing. Additionally, large panel testing, rather than *BAP1* single gene testing, is recommended.

## Introduction

Approximately 12% of unselected UM patients show evidence of a hereditary tumor predisposition syndrome, with an 8-10% chance of detecting a pathogenic variant in a hereditary cancer gene ^1–3^. Risk is higher with early onset disease(age under 30), multiple primary cancers, and/or family history of UM or other cancers ^4,5^.

The National Comprehensive Cancer Network (NCCN) guidelines recommend referral to genetics and genetic testing in UM patients with 1) Early age of diagnosis (<30 years of age); 2) History of other primary cancers in the patient; or a 3) Family or personal history of other cancers known to be associated with a hereditary syndrome ^6^. Only *BAP1, BRCA1, BRCA2, PALB2*, and *MBD4* genes are included as potential candidate genes, and the type of gene testing is not included. The American Society Clinical Oncology (ASCO) guidelines currently recommend single *BAP1* germline testing for patients with UM at risk for hereditary syndrome ^7^. About 1-2% of UM cases have a hereditary *BAP1* pathogenic variant, the most common cause of hereditary UM ^2,8^. Individuals with a *BAP1* pathogenic variant have an increased risk for UM, mesothelioma, renal cell carcinoma, melanoma of the skin, basal cell carcinoma, and other cancers ^9^. We evaluated criteria guidelines and outcomes of genetic panel testing of UM patients referred to The Ohio State University cancer genetics clinic between 05/1/2021 to 9/19/2025.

## Methods

### Retrospective Chart Review

A retrospective chart review was conducted for UM patients seen by a genetic counselor that meet NCCN criteria in The Ohio State University Cancer Genetics Clinic between 5/1/2021-9/19/2025 due to a high risk for hereditary predisposition based on a personal or family history of cancer. This study was conducted under protocols approved by an IRB. During genetic counseling, individuals were offered large panel testing; genes in panel testing changed over the years due to clinical testing company preference (Supplementary Table S1).Two additional patients had prior genetic testing with smaller panels (Supplementary Table S1). Patients had the option to decline clinical genetic testing. All patients were assigned a study ID code, clinical data, including personal and family cancer history, obtained from patient charts. Individuals were assessed via ASCO guidelines for a positive in a gene outside of *BAP1*.

Variant classification was determined from the Ambry Genetics Classifi™ Variant Classification Scheme, a points-based framework which incorporates clinical evidence, molecular impact, and functional assays to determine variant pathogenicity.

### Statistical Analysis

Patients were grouped into three categories, those with P/LP variants, those with VUS, and those who were negative for testing. Significance of difference between various demographic and clinical features between the three groups were assessed using Student’s t-test and Fisher’s exact test (Table 1). Significance of difference between the UM cohort frequency and the GnomAD population frequency was determined using Fisher’s exact test two-sided mid-p value, as previously described ^2^.

**Table 1.** Summary of the Clinical Phenotype of Uveal Melanoma Patients.

|  | P/LP | VUS | Neg | Total<br>(P/LP,<br>VUS, Neg) | <sup>a</sup>p-value |  |  |
| --- | --- | --- | --- | --- | --- | --- | --- |
|  |  |  |  |  | P/LP vs. Neg | P/LP vs. VUS | VUS vs. Neg |
| **Median Age (range)** | 63.5<br>(45-81) | 53<br>(22-76) | 55.5<br>(17-87) | 56 (17-87) | 0.1013 | 0.1574 | 0.7332 |
| **Sex** |  |  |  |  |  |  |  |
| **M** | 2 | 7 | 19 | 28 | 0.2838 | 0.3989 | 1 |
| **F** | 8 | 9 | 25 | 42 |  |  |  |
| **Tumor location** |  |  |  |  |  |  |  |
| **Choroidal** | 8 | 15 | 34 | 57 | 0.4346 | 0.4769 | 0.6999 |
| **Ciliochoroidal** | 1 | 1 | 6 | 8 |  |  |  |
| **Iridociliary** | 0 | 0 | 1 | 1 |  |  |  |
| **Iridociliochoroidal** | 1 | 0 | 0 | 1 |  |  |  |
| **Iris** | 0 | 0 | 3 | 3 |  |  |  |
| **AJCC T Category** |  |  |  |  |  |  |  |
| **1** | 2 | 5 | 17 | 24 | 0.476 | 0.2855 | 0.2031 |
| **2** | 5 | 3 | 15 | 23 |  |  |  |
| **3** | 1 | 4 | 8 | 13 |  |  |  |
| **4** | 2 | 1 | 3 | 6 |  |  |  |
| **Unknown** | 0 | 3 | 1 | 4 |  |  |  |
| **AJCC Stage** |  |  |  |  |  |  |  |
| **I** | 2 | 5 | 17 | 24 | 0.05714 | 0.5577 | 0.1981 |
| **II** | 5 | 7 | 24 | 36 |  |  |  |
| **III** | 3 | 2 | 1 | 6 |  |  |  |
| **IV** | 0 | 0 | 1 | 1 |  |  |  |
| **Unknown** | 0 | 2 | 1 | 3 |  |  |  |
| ***GEP Class*** |  |  |  |  |  |  |  |
| **1A** | 3 | 6 | 13 | 22 |  |  |  |
| **1B** | 0 | 2 | 4 | 6 |  |  |  |
| **2** | 5 | 1 | 9 | 15 | 0.2345 | 0.0783 | 0.6753 |
| **N/A** | 2 | 7 | 18 | 27 |  |  |  |
| ***Primary Treatment*** |  |  |  |  |  |  |  |
| **Brachytherapy** | 8 | 11 | 33 | 52 |  |  |  |
| **Other surgical** | 0 | 0 | 4 | 4 |  |  |  |
| **Other non-surgical** | 0 | 2 | 1 | 3 | 0.7857 | 0.8103 | 0.3312 |
| **Enucleation** | 2 | 3 | 5 | 10 |  |  |  |
| **No Treatment** | 0 | 0 | 1 | 1 |  |  |  |
| ***<sup>b</sup>Metastasis*** |  |  |  |  |  |  |  |
| **Yes** | 0 | 6 | 2 | 8 | 1 | 0.0609 | **0.0019** |
| **No** | 7 | 8 | 40 | 55 |  |  |  |
| ***Personal Cancer History*** |  |  |  |  |  |  |  |
| **Yes** | 6 | 9 | 20 | 35 | 0.4945 | 1 | 0.5634 |
| **No** | 4 | 7 | 24 | 35 |  |  |  |
| ***Family Cancer History*** |  |  |  |  |  |  |  |
| **Yes** | 9 | 16 | 44 | 69 | 0.1852 | 0.3846 | 1 |
| **No** | 1 | 0 | 0 | 1 |  |  |  |

<sup>a</sup>p-value determined by Student's t-test (age) and Fisher's exact test (sex, clinical characteristics, cancer history). <sup>b</sup>Assessed for patients with at least 12-month follow-up in Ophthalmology or who had metastasis present at the time of diagnosis.
Abbreviations: P/LP- pathogenic/Likely pathogenic, VUS- Variant of Uncertain Significance, Neg- Negative, AJCC- American Joint Committee on Cancer, GEP- Gene expression profiling

## Results

Sixty-eight patients were seen for genetic counseling and had large clinical genetic testing, and 2 patients had prior genetic testing with smaller panels (Supplementary Table S1). Of the patients who received testing, 28 were men (40%), 42 (60%) were women, with an age of diagnosis ranging from 17-87 and a mean age of 56 years (Table 1). Most patients had choroidal tumors (81.4%), although tumors with ciliary and iris involvement were also present in this cohort. Most UM patients (74.3%) received primary brachytherapy. Of the patients with at least 12-months follow-up, 12.7% developed metastatic disease. Notably, there was a significant difference between the development of metastasis between patients with VUSs in comparison to patients who with negative results. Out of 14 VUS patients, 6 had metastatic disease progression. Given the small sample size and lack of functional data, interpretations about the underlying pathogenicity of the VUS’s could not be made. Many UM patients had a prior cancer diagnosis (50%), and all but one had family cancer history (98.6%).

Ten individuals tested positive for pathogenic or likely pathogenic (P/LP) alterations in known cancer genes (*BAP1, BRCA1, BRCA2, MBD4, MUTYH, POT1, XRCC2*) (Table 2, Supplementary Table S2). (8/10) of the positives would have been missed if testing was only done for *BAP1*.Two patients were in the same family and positive for the *XRCC2* variant, thus our positive rate in unrelated patients was9/7012.9%) and will be counted as one individual going forward.Importantly, the positive rate for pathogenic variants in an actionable, dominant cancer gene (2 *BAP1, BRCA1, BRCA2, MBD4,2 POT1,)* was 7/70 (10%) Two patients 2/70(2.9%) were noted to be carriers of hereditary cancer syndrome genes, having one copy of a cancer gene with known recessive inheritance (, *MUTYH*, and 2*XRCC2*). Forty-four patients (62.9%, 44/70) tested negative for any germline cancer gene alteration. (Five (7.1%)patients with a P/LP variant also had at least one VUS, and 16 (22.9%) additional patients %) had least one VUS (Table 1; Supplementary Tables S2, S3). *MBD4* was added to the 77-gene panel in November 2024, so 71 patients were not tested for *MBD4*.

**Table 2.** Summary of the Clinical Phenotype of Patients with Pathogenic/Likely Pathogenic Variants.

| Patient ID | Sex | Age Range | Gene Panel<sup>a</sup> | P/LP Variant | VUS | Other Personal Cancer Hx | Family Hx of Cancer | AJCCT Category | AJCC Stage | Treatment Type |
| --- | --- | --- | --- | --- | --- | --- | --- | --- | --- | --- |
| 1 | F | 65-70 | 77a | *BAP1*, c.1526C>A |  | Basal cell carcinoma<br>Endometrial -62 | Mesothelioma, UM, Liver | T2a | IIA | Brachytherapy |
| 2 | F | 60-65 | 71 | *BAP1*, c.1717delC | *MLH1*, c.1874A>G<br>*MSH3*, c.1249C>T | No | Kidney x 3 (26, 62, 50),<br>Melanoma-62, Lung, Prostate, Brain, Colorectal, Cervical, Breast, Stomach | T4b | IIIB | Enucleation |
| 3 | F | 65-70 | 77a | *BRCA1*, c.5109T>G |  | Ovarian-54 | Ovarian, Breast | T2a | IIA | Brachytherapy |
| 4 | F | 40-45 | 77a | *BRCA2*, c.5946delT |  | Breast-44 | Cutaneous melanoma, Lung | T1a | I | Brachytherapy with FNAB |
| 5 | F | 60-65 | 85 | *MBD4*, c.1670T>A | *BRIP1*, c.1489G>C | No | Lung x 2, Melanoma | T2a | IIA | Brachytherapy with FNAB |
| 6 | F | 60-65 | 77a | *POT1*, c.1851_1852del TA | *POT1*, c.670G>A | Lung-79 | Lung x 3, Tonsillar | T2a | IIA | Brachytherapy |
| 7 | F | 75-80 | 71 | *POT1*, c.122dupC |  | Breast-73 | Unknown | T2d | IIIA | Brachytherapy, Enucleation |
| 8 | F | 80-85 | 77a | *MUTYH*, c.734G>A | *BRIP1*, c.2379+4G>A | No | Bladder, Melanoma, Prostate x 3, Basal cell | T3a | IIB | Enucleation |
| 9 | M | 50-55 | 77a | <sup>b</sup>*XRCC2*, c.651_652delTG | *CDKN2A*, c.-2A>T | No | Ovarian- 45<br>Bladder- 92<br>Leukemia- 70<br>Skin | T1a | I | Brachytherapy w/o FNAB |
| 10 | M | 40-45 | 77a | <sup>b</sup>*XRCC2*, c.651_652delTG | *CDKN2A*, c.-2A>T | Thyroid-40<br>Basal cell-76 |  | T4a | IIIA | Brachytherapy, Laser, Enucleation |

<sup>a</sup>Gene panels 77a, 71 did not include *MBD4*; Gene panels 71, 85 did not include *XRCC2* (see Supplementary Table 1). <sup>b</sup>These individuals are related.
Abbreviations: Age, Age at diagnosis; P/LP, pathogenic/Likely pathogenic; VUS, Variant of Uncertain Significance; FNAB, fine needle aspiration biopsy

We compared the frequency of P/LP variants in our cohort to that of a control population (gnomAD v4.0, non-Finnish European group) (Table 3). Three genes (*BAP1, POT1, XRCC2*) showed a significantly higher frequency of P/LP variants in our cohort relative to the control population. Notably, a P/LP variant in one gene (*XRCC2*) has not been previously reported in UM patients.

**Table 3.** Frequencies of P/LP variants in UM cohort and public database.

| Gene | UM cohort P/LP carrier frequency | <sup>a</sup>Population P/LP variant frequency | Odds Ratio (95% CI) | <sup>b</sup>P-value | Published UM cases with P/LP variants |
| --- | --- | --- | --- | --- | --- |
| *BAP1* | 2/70 (2.9%) | 77/590031 (0.013%) | 218.9 (35.5 - 781.1) | **4.509e-05** | >50 reported cases |
| *BRCA1* | 1/69 (1.4%) | 733/590031 (0.124%) | 11.7 (0.6 – 59.1) | 0.08697 | 4<sup>2,10-13</sup> |
| *BRCA2* | 1/70 (1.3%) | 1913/590031 (0.324%) | 4.4 (0.2 - 22.2) | 0.22850 | 8<sup>13-16</sup> |
| *MBD4* | 1/5 (20%) | 949/590031 (0.161%) | N/A | N/A | 24<sup>12,16-21</sup> |
| *MUTYH* | 1/68 (1.5%) | 574/590031 (0.097%) | 15.1 (0.7 – 76.7) | 0.06719 | N/A |
| *POT1* | 1/69 (1.4%) | 269/590031 (0.046%) | 63.5 (10.4 – 219.8) | **0.00051** | 3<sup>3,22</sup> |
| *XRCC2* | <sup>c</sup>1/54 (1.9%) | 397/590031 (0.067%) | 27.5 (1.3 – 140.8) | **0.03709** | N/A |

<sup>a</sup> The gnomAD version 4.1.0 non-Finnish European cohort frequency was estimated by the number of variant alleles reported as P/LP variants and null variants not reported as benign or of uncertain significance. <sup>b</sup> P-value determined by Fisher's exact test with two-sided mid-p value. <sup>c</sup> Frequency of XRCC2 mutations excluded related individuals.
Abbreviations: P/LP, pathogenic/Likely pathogenic; UM, Uveal Melanoma; CI, Confidence Interval; N/A, No Answer

## Discussion

This is the first study to assess outcomes of NCCN guidelines for testing high-risk UM patients. Seven of the P/LP variants were in actionable/potentially actionable genes with available management guidelines (*BAP1 (2), BRCA1, BRCA2*,, *MBD4, POT1 (2)*,) highlighting the importance of referring UM patients with a strong personal and/or family history of cancer to genetic counseling and testing ^6^. Seven individuals had a pathogenic variant in a gene that has been reported previously in UM patients (*BAP1 (2), BRCA1, BRCA2, MBD4, POT1 (2)*,) ^2,3,10–22^. Currently, the NCCN UM genetic testing guidelines list five genes *BAP1, BRCA1, BRCA2, PALB2*, and *MBD4* to be associated with genetic predisposition to UM while the ASCO guidelines recommend testing for only a single gene, *BAP1* ^6,7^. Our findings reinforce the utility of large multigene panel genetic testing, rather than small panel or single gene, for UM patients with features suggestive of hereditary cancer risk.

This study supports our earlier finding of phenotypic and locus heterogeneity in patients with evidence of UM-related tumor predisposition, consistent with findings from other groups ^2^. A recent French population-based study, Le Ven et al., analyzed 381 patients with UM and identified 21 pathogenic variants across *BAP1* (3), *MBD4* (7), *BRCA2* (3), *MLH1* (1), *MSH2* (1), *MSH6* (2), and *PMS2* (4), supporting the possibility that individuals with Lynch syndrome are also at risk for UM ^16^. In addition, a Finnish germline study of 270 unselected UM patients-excluding those with known pathogenic variants in *BAP1* or *MBD4-* identified single P/LP variants in *BRCA1*, BLM and *MET*. ^10^. This data further broadens the spectrum of cancer-predisposition genes associated with UM and supports use of comprehensive germline testing strategies.

Across our current and prior studies, we have identified germline P/LP variants in at least 10 additional cancer genes have been reported in UM patients^2,3,10–22^. Identification of germline P/LP variants in *BRCA1, BRCA2* and in this study provide additional, although limited, evidence for the potential association of these genes with predisposition to UM. Since the majority of UM are treated with radiation and tumor tissue is rarely available, we were unable to assess biallelic inactivation (second hit) in the tumor.

The contribution of other genes, including, *MUTYH*, and *XRCC2*, to UM risk remains uncertain and warrants further investigation. *MUTYH* and *XRCC2* are associated with autosomal recessive cancer syndromes—MUTYH-associated polyposis and Fanconi anemia, respectively—where biallelic inactivation is typically required for disease manifestation. However, single P/LP variants in both genes have been reported in patients with various malignancies, raising the possibility that single pathogenic variants may confer increased cancer risk^25^. Additional studies are needed to clarify whether such variants contribute to UM.

Although the contribution of numerous genes to UM risk remains uncertain, our results expand the possible germline landscape of UM and highlight its genetic heterogeneity. Future studies integrating germline and somatic sequencing with functional assays will be critical to establish causality, refine risk estimates, and inform evidence-based updates to genetic testing guidelines. As large-scale genomic datasets and collaborative registries continue to grow, a clearer understanding of UM predisposition genes will emerge—advancing both precision risk assessment and mechanistic insight into this rare malignancy.

## Conclusions

Current NCCN guidelines are appropriate for identification of high-risk UM patients for testing and using a large genetic panel rather than a single gene or limited genes panel is highly recommended.

## Data Availability

The data presented in this study are available on request from the corresponding author.

## Abbreviations

(UM): Uveal melanoma
(NCCN): National Comprehensive Cancer Network
(ASCO): The American Society Clinical Oncology pathogenic or likely pathogenic (P/LP)
(VUS): variance of uncertain significance

## Author Contributions

Conceptualization, LB, CC, MAR; Methodology, LB, IG, CC, MAR; Analysis, LB, JM, OT; Data Curation, LB, IG, KR; Writing-Original Draft Preparation, LB, IG, MAR; Writing-Review and Editing, LB, IG, KR, JM, OT

## Supplementary Data

Supplementary Table 1: Genetic Testing Panels

Supplementary Table 2: P/LP and VUS Protein Consequence & Reference ID Summary

Supplementary Table 3: Summary of the Clinical Phenotype of Patients with Variants of Uncertain Significance

