## Supplementary Tables 1 & 2 for "Multigene Panel Testing Outcomes in Patients with Uveal Melanoma: Implementation of National Guidelines"

**Supplementary Table 1: Genetic Testing Panels**

| ***Ambry CancerNext-Expanded*** | | | | ***Invitae*** | |
| --- | --- | --- | --- | --- | --- |
| **77a (July 2020-February 2024** | **71 (February 2024- November 2024** | **76/77b (standard panel, November 2025-)** | **85 (custom panel, November 2025-)** | **16** | **9** |
| *AIP*  *ALK*  *APC*  *ATM*  *AXIN2*  *BAP1*  *BARD1*  *BLM*  *BMPR1A*  *BRCA1*  *BRCA2*  *BRIP1*  *CDC73*  *CDH1*  *CDK4*  *CDKN1B*  *CDKN2A*  *CHEK2*  *CTNNA1*  *DICER1*  *EGFR*  *EGLN1*  *EPCAM*  *FANCC*  *FH*  *FLCN*  *GALNT12*  *GREM1*  *HOXB13*  *KIF1B*  *KIT*  *LZTR1*  *MAX*  *MEN1*  *MET*  *MITF*  *MLH1*  *MSH2*  *MSH3*  *MSH6*  *MUTYH*  *NBN*  *NF1*  *NF2*  *NTHL1*  *PALB2*  *PDGFRA*  *PHOX2B*  *PMS2*  *POLD1*  *POLE*  *POT1*  *PRKAR1A*  *PTCH1*  *PTEN*  *RAD51C*  *RAD51D*  *RB1*  *RECQL*  *RET*  *SDHA*  *SDHAF2*  *SDHB*  *SDHC*  *SDHD*  *SMAD4*  *SMARCA4*  *SMARCB1*  *SMARCE1*  *STK11*  *SUFU*  *TMEM127*  *TP53*  *TSC1*  *TSC2*  *VHL*  *XRCC2* | *AIP*  *ALK*  *APC*  *ATM*  *AXIN2*  *BAP1*  *BARD1*  *BMPR1A*  *BRCA1*  *BRCA2*  *BRIP1*  *CDC73*  *CDH1*  *CDK4*  *CDKN1B*  *CDKN2A*  *CHEK2*  *CTNNA1*  *DICER1*  *EGFR*  *EGLN1*  *EPCAM*  *FH*  *FLCN*  *GREM1*  *HOXB13*  *KIF1B*  *KIT*  *LZTR1*  *MAX*  *MEN1*  *MET*  *MITF*  *MLH1*  *MSH2*  *MSH3*  *MSH6*  *MUTYH*  *NF1*  *NF2*  *NTHL1*  *PALB2*  *PDGFRA*  *PHOX2B*  *PMS2*  *POLD1*  *POLE*  *POT1*  *PRKAR1A*  *PTCH1*  *PTEN*  *RAD51C*  *RAD51D*  *RB1*  *RET*  *SDHA*  *SDHAF2*  *SDHB*  *SDHC*  *SDHD*  *SMAD4*  *SMARCA4*  *SMARCB1*  *SMARCE1*  *STK11*  *SUFU*  *TMEM127*  *TP53*  *TSC1*  *TSC2*  *VHL* | *AIP*  *ALK*  *APC*  *ATM*  *AXIN2*  *BAP1*  *BARD1*  *BMPR1A*  *BRCA1*  *BRCA2*  *BRIP1*  *CDC73*  *CDH1*  *CDK4*  *CDKN1B*  *CDKN2A*  *CEBPA*  *CHEK2*  *CTNNA1*  *DDX41*  *DICER1*  *EGFR*  *EPCAM*  *ETV6*  *FH*  *FLCN*  *GATA2*  *GREM1*  *HOXB13*  *KIT*  *LZTR1*  *MAX*  *MBD4*  *MEN1*  *MET*  *MITF*  *MLH1*  *MSH2*  *MSH3*  *MSH6*  *MUTYH*  *NF1*  *NF2*  *NTHL1*  *PALB2*  *PDGFRA*  *PHOX2B*  *PMS2*  *POLD1*  *POLE*  *POT1*  *PRKAR1A*  *PTCH1*  *PTEN*  *RAD51C*  *RAD51D*  *RB1*  *RET*  *^a^RPS20*  *RUNX1*  *SDHA*  *SDHAF2*  *SDHB*  *SDHC*  *SDHD*  *SMAD4*  *SMARCA4*  *SMARCB1*  *SMARCE1*  *STK11*  *SUFU*  *TMEM127*  *TP53*  *TSC1*  *TSC2*  *VHL*  *WT1* | *AIP*  *ALK*  *APC*  *ATM*  *AXIN2*  *BAP1*  *BARD1*  *BMPR1A*  *BRCA1*  *BRCA2*  *BRIP1*  *CDC73*  *CDH1*  *CDK4*  *CDKN1B*  *CDKN2A*  *CEBPA*  *CHEK2*  *CTNNA1*  *DDX41*  *DICER1*  *EGFR*  *EPCAM*  *ETV6*  *FH*  *FLCN*  *GATA2*  *GREM1*  *HOXB13*  *KIT*  *LZTR1*  *MAX*  *MBD4*  *MEN1*  *MET*  *MITF*  *MLH1*  *MSH2*  *MSH3*  *MSH6*  *MUTYH*  *NF1*  *NF2*  *NTHL1*  *PALB2*  *PDGFRA*  *PHOX2B*  *PMS2*  *POLD1*  *POLE*  *POT1*  *PRKAR1A*  *PTCH1*  *PTEN*  *RAD51C*  *RAD51D*  *RB1*  *RET*  *RPS20*  *RUNX1*  *SDHA*  *SDHAF2*  *SDHB*  *SDHC*  *SDHD*  *SMAD4*  *SMARCA4*  *SMARCB1*  *SMARCE1*  *STK11*  *SUFU*  *TMEM127*  *TP53*  *TSC1*  *TSC2*  *VHL*  *WT1*  *ATRIP*  *EGLN1*  *KIF1B*  *MLH3*  *PALLD*  *RAD51B*  *RNF43*  *TERT* | *ATM*  *BAP1*  *BARD1*  *BRCA1*  *BRCA2*  *BRIP1*  *CDH1*  *CHEK2*  *MSH2*  *NBN*  *NF1*  *PALB2*  *PTEN*  *RAD50*  *STK11*  *TP53* | *BAP1*  *BRCA2*  *CDK4*  *CDKN2A*  *POT1*  *PTEN*  *RB1*  *TP53*  *MITF* |

^a^RPS20 added to panel in April 2025.

**Supplementary Table 2: P/LP and VUS Protein Consequence & Reference ID Summary**

| **Patient ID** | ^a^**ClinVar Classification** | **Gene** | **Variant name** | **Predicted Protein Consequence** | **Gene Reference ID** |
| --- | --- | --- | --- | --- | --- |
| 1 | P | *BAP1* | c.1526C>A | p.Ser509Ter | NM_004656.4 |
| 2 | P | *BAP1* | c.1717delC | p.Leu573Trpfs*3 | NM_004656.4 |
|  | VUS/LB | *MLH1* | c.1874A>G | p.Tyr625Cys | NM_000249.4 |
|  | VUS | *MSH3* | c.1249C>T | p.Arg417Trp | NM_002439.5 |
| 3 | P | *BRCA1* | c.5109T>G | p.Tyr1703Ter | NM_007294.4 |
| 4 | P | *BRCA2* | c.5946delT | p.Ser1982fs | NM_000059.4 |
| 5 | P/LP | *MBD4* | c.1670T>A | p.Leu557Ter | NM_001276270.2 |
|  | VUS | *BRIP1* | c.1489G>C | p.Val497Leu | NM_032043.3 |
| 6 | LP | *POT1* | c.1851_1852delTA | p.Asp617Glufs*9 | NM_015450.3 |
|  | VUS | *POT1* | c.670G>A | p.Asp224Asn | NM_015450.3 |
| 7 | LP | *POT1* | c.122dupC | p.Asp42Ter | NM_015450.3 |
| 8 | P | *MUTYH* | c.734G>A | p.Arg245His | NM_001048174.2 |
|  | VUS | *BRIP1* | c.2379+4G>A | Unknown | NM_032043.3 |
| 9,10 | LP | *XRCC2* | c.651_652delTG | p.Cys217Ter | NM_005431.2 |
|  | VUS/LB | *CDKN2A* | c.-2A>T | Unknown | NM_058195.4 |
| 11 | VUS/LB | *ATM* | c.6759A>C | p.Lys2253Asn | NM_000051.4 |
| 12 | VUS | *ATM* | c.1225C>G | p.Leu409Val | NM_000051.4 |
| 13 | VUS | *AXIN2* | c.1058C>T | p.Pro353Leu | NM_004655.4 |
| 14 | VUS | *BAP1* | c.1152C>G | p.Ser384Arg | NM_004656.4 |
|  | VUS | *NTHL1* | c. 931G>C | p.Gly311Arg | NM_002528.7 |
| 15 | VUS/LB | *BARD1* | c.1360C>G | p.Pro454Ala | NM_000465.4 |
| 16 | VUS | *BLM* | c.270G>A | p.Ala914Thr | NM_000057.4 |
| 17 | VUS | *BRIP1* | c.2594G>A | p.Arg865Gln | NM_032043.3 |
| 18 | VUS | *CDH1* | c.2629G>A | p.Gly877Arg | NM_004360.5 |
| 19 | VUS | *CHEK2* | c.1141A>G | p.Met381Val | NM_007194.4 |
| 20 | VUS | *CTNNA1* | c.911C>T | p.Ser304Phe | NM_001903.5 |
| 21 | VUS | *MET* | c.1510G>T | p.Val504Phe | NM_000245.4 |
| 22 | VUS | *PMS2* | c.1126C>G | p.Pro376Ala | NM_000535.7 |
| 23 | VUS | *RECQL* | c.1025G>A | p.Gly342Asp | NM_002907.4 |
| 24 | VUS | *SDHB* | c.457A>G | p.Ile153Val | NM_003000.3 |
| 25 | VUS | *TMEM127* | c.526A>G | p.Ile188Val | NM_017849.4 |
| 26 | VUS/B/LB | *TSC2* | c.3818C>T | p.Ala1273Val | NM_000548.5 |

^a^VUS classification was determined by Ambry Variant Classification Scheme 2023, but ClinVar VUS or conflicting classifications are summarized in this column.

Abbreviations: B, Benign; LB, Likely Benign; P, Pathogenic; LP, Likely Pathogenic; VUS, Variant of Uncertain Significance

**Supplementary Table 3:** **Summary of the Clinical Phenotype of Patients with Variants of Uncertain Significance**

| **Patient ID** | **Sex** | **Age of Dx** | **Gene Panel** | **Gene** | **Other Personal Cancer Hx** | **Family Hx of Cancer** | **AJCCT**  **Category** | **AJCC Stage** | **Treatment type** |
| --- | --- | --- | --- | --- | --- | --- | --- | --- | --- |
| 11 | F | 50-55 | 16 | *ATM,* c.6759A>C | Breast | Colorectal, Skin, Endometrial | T3b | IIIA | Brachytherapy with FNAB |
| 12 | M | 30-35 | 77a | *ATM*, c.1225C>G | None | Melanoma | T4b | IIIB | Enucleation |
| 13 | F | 45-50 | 77a | *AXIN2,* c.1058C>T | None | Melanoma, Ovarian | T2a | IIa | Brachytherapy with FNAB, TTT |
| 14 | M | 65-70 | 77a | *BAP1,* c.1152C>G  *NTHL1,* c.931G>C | None | Kidney, Basal Cell Carcinoma, Prostate | T1a | I | Brachytherapy with FNAB |
| 15 | F | 65-70 | 71 | *BARD1*  c.1360C>G | Basal cell carcinoma | Melanoma  Leukemia-CLL  Prostate  Breast- 81,  Lung- 51,  Colon- 77  Leukemia- 88  Basal cell carcinoma | T3b | IIB | Brachytherapy |
| 16 | F | 60-65 | 77a | ^a^*BLM,* c.270G>A | None | Skin, Prostate | T1a | I | Brachytherapy with FNAB |
| 17 | F | 40-45 | 77a | *BRIP1,* c.2594G>A | None | Breast | Unknown | IIA | Enucleation |
| 18 | M | 75-80 | 77a | *CDH1,* c.2629G>A | Melanoma | None | T1a | I | TTT |
| 19 | M | 50-55 | 77a | *CHEK2,* c.1141A>G | Basal cell carcinoma | None | Unknown | Unknown | Brachytherapy |
| 20 | F | 50-55 | 77a | *CTNNA1,* c.911C>T | None | Prostate (metastatic) | T3a | IIB | Brachytherapy, enucleation |
| 21 | F | 40-45 | 77a | *MET,* c.1510G>T | None | None | Unknown | Unknown | Brachytherapy, enucleation |
| 22 | M | 70-75 | 77a | *PMS2,* c.1126C>G | None | Breast | T1a | I | Brachytherapy with FNAB |
| 23 | F | 70-75 | 77a | *RECQL,* c.1025G>A | Melanoma | None | T2a | IIA | Vitrectomy, FNAB, laser |
| 24 | M | 65-70 | 77a | *SDHB,* c.457A>G | None | Pancreatic | T1a | I | Brachytherapy with FNAB |
| 25 | F | 20-22 | 71 | *TMEM127*  c.526A>G | None | Breast- 35  Esophageal | T3a | IIB | Enucleation |
| 26 | M | 45-50 | 76 | *TSC2*  c.3818C>T | None | Basal cell  Lung  Throat | T2a | IIA | Brachytherapy |

^a^Pathogenic variant associated with recessive condition. ^b^The patient reported family history of cancer, but the types are unknown.

Abbreviations: Dx, diagnosis; Hx, history; AJCC, American Join Committee on Cancer; ASCO, American Society of Clinical Oncology; NCCN, National Comprehensive Cancer Network; FNAB, Fine-needle aspiration biopsy; TTT, transpupillary thermotherapy
